# Neuropsychological and behavioral profiles of patients with chronic trigeminal neuralgia

**DOI:** 10.1101/2024.10.30.24316421

**Authors:** Anton Pashkov, Elena Filimonova, Azniv Martirosyan, Galina Moisak, Jamil Rzaev

## Abstract

Patients suffering from chronic pain are known to exhibit distinctive personality traits and impaired neuropsychological performance across various cognitive domains. However, there is currently a lack of comprehensive evidence regarding cognitive and behavioral functioning patterns in patients with trigeminal neuralgia. In this study, we aimed to thoroughly characterize a range of psychological and neuropsychological variables in a sample of 80 patients and 34 healthy controls, and to assess their relationship with pain intensity and duration. Our findings revealed that patients with trigeminal pain scored significantly higher on measures of anxiety, depression, perceived stress, alexithymia, pain catastrophizing, harm avoidance and lower on Self-transcendence subscale compared to healthy controls. Additionally, these patients demonstrated lower performance scores on tasks assessing working memory and verbal fluency. These findings may provide valuable insights for the development of personalized treatment plans for patients with trigeminal neuralgia, specifically targeting their unique personality traits and cognitive impairments.

## Introduction

Diagnostics and treatment of chronic pain syndromes pose a significant challenge for healthcare systems globally (Cohen et al., 2021). One specific type of chronic pain is trigeminal neuralgia, which is characterized by intermittent episodes of sharp, electric shock-like facial pain. This condition significantly impacts the quality of life of those affected and hinders basic daily activities such as oral hygiene, eating, or speaking (Bendtsen et al., 2020). The primary approach to managing pain for these patients typically involves the regular use of anti-epileptic drugs (AEDs) and/or antidepressants. While these medications can be useful, they can also cause adverse effects and may lose their effectiveness over time in relieving pain, prompting many patients to seek alternative treatments. Since compression of the trigeminal nerve at the root entry zone by adjacent vessels is believed to be the main cause of trigeminal pain in such cases, surgical intervention often emerges as an effective, generally safe, and accessible alternative for individuals who do not respond well to pharmacological treatments (Xu et al., 2021).

It is noteworthy that mental health issues are prevalent among patients with trigeminal neuralgia, including those undergoing surgical treatment, yet they are often neglected in pain management protocols (Darnall et al., 2016). The current focus in identifying reliable predictors of treatment outcomes primarily lies with MRI-derived biomarkers, which are considered more reliable and promising than those obtained from psychological or psychiatric assessments. However, recent studies have shown the importance of incorporating patients’ psychological status and neuropsychological performance scores into treatment planning (Attal et al., 2014; Driscoll et al., 2021). In patients with chronic pain who did not undergo surgery, psychological factors were found to be strong predictors of functional treatment outcomes at 6 and 12 months after treatment (George and Beneciuk, 2015; Trinderup et al., 2018). Furthermore, comorbid anxiety and depression symptoms, along with beliefs about the negative impact of pain on health, its perceived uncontrollable nature, and anticipated persistence, have been shown to directly influence pain perception. Impaired executive functions have also been demonstrated to play a role in this process (Simons et al., 2014; Cormier et al., 2016; Meints and Edwards, 2018).

Earlier studies involving diverse groups of patients with various chronic pain conditions showed that these individuals had higher levels of harm avoidance and lower levels of self-directedness, as assessed by Cloninger’s temperament and character inventory (Knaster et al., 2012; Conrad et al., 2013). Additionally, it has been observed that individuals with a long-standing history of chronic pain exhibited increased levels of alexithymia (Aaron et al., 2019). Previous studies have demonstrated that a higher prevalence of alexithymia in the general population is associated with an increased risk of developing chronic pain (Makino et al., 2013). Moreover, psychological constructs such as fear of pain and catastrophizing beliefs are widely acknowledged as important factors in understanding the mechanisms of chronic pain, as they are commonly identified as key psychological variables that directly affect all aspects of pain behavior (Turk et al., 2010; Zale and Ditre, 2015).

Furthermore, drawing a parallel to the classification of type A and type B personalities, which have been established to define distinct personality traits associated with an increased risk of cardiovascular diseases, some researchers advocate for the introduction of a concept called «pain personality». This concept aims to encompass the diverse behavioral characteristics observed in chronic pain patients, potentially facilitating a more comprehensive evaluation and the ability to predict functional treatment outcomes and resistance to treatment (Gustin et al., 2015; Naylor et al., 2017).

Emerging evidence from various chronic pain syndromes suggests that certain conditions may share similar behavioral profiles and show declines in performance across similar cognitive domains (Higgins et al., 2018). Additionally, recent findings indicate that patients with chronic pain experience higher rates of cognitive decline compared to age– and gender-matched pain-free cohorts, particularly affecting verbal memory, semantic fluency, and temporal orientation abilities (Rong et al., 2021).

The relationship between pain severity and cognitive functions is two-way, with these two factors intricately intertwined and influencing each other (Berryman et al., 2014). Different high-level neuropsychological subdomains have been implicated in the development of chronic pain, potentially contributing to the transition from acute to chronic pain states (Edwards et al., 2016; Mazza et al., 2018). Studies have demonstrated that lower scores on attention and memory assessments correlate with increased pain intensity and interference in individuals with chronic pain (Liu et al., 2013, Baker et al., 2017). Importantly, this association between neuropsychological performance measures and pain intensity extends beyond patient populations to include non-clinical samples as well (Nakae et al., 2013). Moreover, interventions aimed at specific cognitive or behavioral factors have shown effectiveness not only in restoring optimal psychological functioning but also in relieving pain, suggesting a causal relationship between these variables that goes beyond mere correlations (Yoshino et al., 2018).

At the same time, in stark contrast to the wealth of knowledge accumulated in studying other chronic pain conditions, there is currently a noticeable lack of research dedicated to exploring the unique behavioral traits and neurocognitive performance in patients with trigeminal neuralgia. To date, most published studies on individuals with orofacial pain have primarily focused on assessing levels of anxiety, perceived stress, pain catastrophizing, and depression (Gustin et al., 2011; Jasim et al., 2014; Cundiff-O’Sullivan et al., 2023). Some researchers have also highlighted the potential risk of developing post-traumatic stress disorder in trigeminal neuralgia patients (Jiang, 2018). While undoubtedly valuable, the existing data only begin to explore patients’ pain-related behaviors, overlooking a wide range of potentially clinically relevant variables.

The goal of this study was to conduct a comprehensive analysis of the psychological and neurocognitive characteristics of patients diagnosed with primary trigeminal neuralgia who underwent surgical intervention at the neurosurgical hospital. We also aimed to evaluate the predictive power of these psychological and neuropsychological variables in determining the level of pain intensity reported by patients.

## Methods

### Participants and study design

From January 2022 to January 2024, a prospective case-control study was carried out at our hospital as a part of an ongoing scientific project. The primary objective of this project was to comprehensively explore neuroimaging, neuropsychological, and blood biomarkers associated with chronic facial pain in patients with primary trigeminal neuralgia, while also identifying potential predictors of surgical outcomes. In this article, we limit ourselves to presenting the results pertaining to the investigation of psychological and neuropsychological variables. Eighty patients with trigeminal neuralgia and 34 healthy age and sex-matched healthy subjects were enrolled and took part in this study. The study comprised of patients diagnosed with primary trigeminal neuralgia who were referred for surgical treatment due to either the inefficacy or severe side effects of their pharmacological therapy (n = 73, age range: 27-83, 62% female). Additionally, a small subset of patients (n = 7, age range: 49-70, 71% female) who received outpatient care and conservative management at our Center were included as well. Sex and age-matched healthy controls (HC, n = 34, age range: 38-75, 59% female) were recruited through local advertisements targeting hospital personnel and their relatives. Evaluation of changes in the cognitive and behavioral status of patients was carried out six months after surgery. Patients were invited to our neurosurgical center again for an MRI, blood test, neurological examination and psychological/neuropsychological assessment.

The inclusion criteria for patients in this study were as follows: experiencing trigeminal nerve pain, no contraindications for MRI, absence of intracranial and facial anomalies, no other neurological or psychiatric disorders, MMSE score > 24, right-handedness. All patients with classical and idiopathic trigeminal neuralgia met the diagnostic criteria outlined in The International Classification of Headache Disorders, 3rd edition (Headache Classification Committee of the International Headache Society, 2018). We specifically enrolled patients from these groups and excluded cases of trigeminal neuralgia secondary to multiple sclerosis, intracranial tumors, and other craniofacial pathological conditions to minimize confounding factors. All patients provided written informed consent to participate in the study. Patient consents were obtained according to the Declaration of Helsinki. Ethics approval for the research protocol was given by local ethic committee of Federal Center of Neurosurgery dated 25.05.2021 (protocol # 7).

### Psychological assessment

Psychological assessment of patients was carried out on the day of hospital admission. For this purpose, we used a number of psychological questionnaires and scales to measure several key psychological constructs and personality traits. The forty-item Spielberger State-Trait Anxiety Inventory was utilized to evaluate both the patient’s current symptoms of anxiety and their long-term tendency to experience anxiety in different situations over an extended period of time (Spielberger, et al., 1970). Perceived stress scale (PSS-10) was included in the diagnostic battery in order to assess the level at which individuals perceive situations in their life as stressful (Cohen et al., 1983). Another relevant concept is an intolerance to uncertainty, which is considered a cognitive vulnerability factor that is shared across different anxiety disorders. In this study, it was measured with the eponymous 22-item scale developed by D. McLain (McLain, 1993). Toronto alexithymia scale (TAS-20) was administered to quantify individual difficulties in identifying and describing their own feelings (Bagby et al., 1994). Pain catastrophizing scale, consisting of 13 statements, was additionally given to all patients (Sullivan et al., 1995). Catastrophizing is often characterized as an amplified negative mindset towards unpleasant stimuli and has a significant influence on the perception and management of pain. Finally, we offered patients to fill in Temperament and Character inventory (TCI-140) in order to gain further information regarding their personality traits since results of previous studies were indicative of noticeable differences between patients with chronic pain and healthy participants on some of these traits (Cloninger, 1994). The subscales of the TCI-140 were analyzed separately, as there is no overall score for the entire test. Healthy volunteers completed the same scales and questionnaires as the patients during their one-day visit to the hospital.

### Neurocognitive testing

At the first day of hospital stay we also administered a battery of neuropsychological tests to evaluate executive functions, such as attention, behavioral flexibility, working memory as well as short-term and long-term memory for both verbal and nonverbal stimuli. The choice of tasks was informed by an extensive review of scientific literature, aiming to identify previous studies that highlighted cognitive impairments in individuals with chronic pain. The final battery comprised five tests: Mini Mental State Examination Scale (MMSE), Rey-Osterrieth complex figure test (ROCF), trail making test (TMT), digit span task, and verbal fluency task. Additionally, 10-item Edinburgh handedness inventory was used to objectively determine a subject’s handedness in daily living activities. Patients were assessed six months after the surgery using the same tests except for MMSE.

The MMSE scale is designed to serve as a concise and easily administered tool for cognitive screening, allowing for the prompt detection of severe cognitive impairments (Folstein et al., 1975). In the current study, patients who scored less than 24 points out of 30 on this test were excluded from further analysis. The Rey–Osterrieth complex figure task was conducted in three parts (Rey and Osterrieth, 1941). In the first part, participants were asked to copy the given complex figure as accurately as possible. The second part was an immediate recall phase, where the patient’s goal was to draw the previously shown figure from memory. The same task given to patients twenty minutes later constituted a delayed recall phase. Final scoring was performed according to the scheme introduced by Petilli and colleagues, which involved dividing the complex figure into 48 basic elements and awarding 1 point for each correctly reproduced element (Petilli et al., 2021).

The Trail Making Test, consisting of two main parts, is one of the most widely used and reliable tools for evaluating visual attention and cognitive flexibility in clinical practice (Reitan, 1956). In part A, the subject is given instructions to connect a series of 25 dots in ascending numerical order with the goal of completing the task as rapidly as possible while ensuring accuracy. In turn, part B places an extra emphasis on attention flexibility, complicating the task by adding the requirement of switching between digits and letters in the following way: 1-A, 2-B, 3-C, and so on. Since direct TMT scores, that is, completion times for parts A and B, are strongly influenced by age, education, and other sociocultural factors, this study used the ratio score (B/A) as an index of attention performance instead (Christidi et al., 2015).

The Digit Span is a cognitive assessment tool used to measure verbal short-term and working memory. It consists of two formats: the Forward Digit Span (WAIS-F) and the Backward Digit Span (WAIS-B), both originally subscales of the Wechsler Adult Intelligence Scale (Wechsler, 2009). In this verbal task, auditory stimuli are presented to participants, who then have to verbally repeat the series of digits either in the same order as presented (forward span) or in reverse order (backward span). The WAIS-F assesses verbal working memory and attention, while the WAIS-B additionally evaluates cognitive control and executive functions. Verbal fluency tests are another commonly used measure to assess executive dysfunction. They typically involve generating words within a specific time frame. Two main types of these tasks were employed in this study: phonological verbal fluency task, where words starting with a given letter are produced and categorical verbal fluency task, where words within a specific semantic category are generated. The final score for each task was the number of words participants could generate in 60 seconds. These tasks measure the functioning of the semantic system, which is responsible for storing words and their associations. Additionally, they evaluate the effectiveness of retrieval strategies and the individual’s ability for self-monitoring and inhibiting inappropriate responses.

### Pain measurement

The severity of a patient’s pain upon admission to the hospital and during the 6-month follow-up was evaluated using the Brief Pain Inventory (BPI; Cleeland, 1991). The BPI employs an 11-point numerical rating scale (0-10), prompting patients to rate the minimum, maximum, and average intensity of their pain over the past week. Additionally, the BPI evaluates the impact of pain on various aspects of a patient’s daily life and assists in identifying clinically significant characteristics of pain, such as tingling, burning, or numbness sensations.

### Surgical complications and outcomes

Postoperative complications were assessed using a scale developed by Theodosopoulos and colleagues (Theodosopoulos et al., 2012). All complications were divided into minor and major. For example, minor complications included allergic reactions, deep vein thrombosis, prolonged inpatient stay, wound hematoma, or numbness that occurred after microvascular decompression. Major complications included unplanned reoperation, cerebrospinal fluid leakage, meningitis, pneumonia, and new-onset seizure disorder.

The outcome of the surgery (favorable or unfavorable) was determined based on the degree of pain reduction after the intervention. We established a strict threshold and only considered outcomes favorable if the patient experienced complete pain relief (100% reduction in pain) six months after undergoing surgery. In other cases, the outcome was considered unfavorable.

### Statistical analysis

For continuous variables, we reported the mean ± standard deviation if the distribution was normal, and the median (interquartile range, IQR) if it deviated from normality. The Shapiro-Wilk test was used to assess the normality of the distribution. Categorical variables were presented as frequency and percentage. The analysis of categorical data was performed using Pearson’s chi-squared test with Yates’ continuity correction. For normally distributed data, the t-test with Welch correction was used. In cases where the normality assumption was violated, the Mann-Whitney test was employed. We also checked for homogeneity of variances in the experimental groups using the Levene test. Moreover, for those neuropsychological variables that showed statistically significant differences between the groups of patients and healthy volunteers, we additionally performed ANCOVA to ensure that the results were not due to the influence of confounding variables (educational attainment, depression and anxiety symptoms, as well as pain catastrophizing scores). For those models, we also reported effect size estimates (partial eta-squared values) and corresponding 95% confidence intervals. The raw neuropsychological data was not transformed into z-scores due to the lack of culture-specific normative neuropsychological data. To examine the relationship between clinical and behavioral variables Spearman partial correlations were utilized (accounting for the effect of age). Wilcoxon rank-sum test was utilized to compare the median values of psychological and neuropsychological variables before and after surgery. To estimate the strength of the relationship between two variables, we calculated the effect size measured as d-Cohen (for t-test). For the values obtained using Wilcoxon and Mann–Whitney tests, we used Glass’ and matched pairs rank biserial correlation coefficients (r_rb_) as estimates of effect size.

Additionally, to predict pain intensity level (taken as mean BPI score) we leveraged multiple linear regression approach. All assumptions (normal distribution and independence of residuals, homoscedasticity, multicollinearity, influential data points) were checked and reported in the Supplemental file. For the regression analysis, all variables in the dataset with more than 30% missing values were removed. Variables with less than 5% missing values were imputed using median values, while those with 5–30% missing data were imputed using nonparametric Random Forest approach. Four methods were used to select predictors for the regression model in order to avoid the influence of a particular method on the final result, namely: Boruta (random forest classification algorithm), MARS (Multivariate Adaptive Regression Splines), stepwise regression approach, and an algorithm based on relative importance (RIA). Leave-one-out cross-validation (LOOCV) was used in order to assess the model’s performance on new data. Root mean squared error (RMSE) and mean absolute error (MAE) were subsequently calculated. A p-value of 0.05 was considered the threshold for determining statistical significance. To account for multiple comparisons, we applied False Discovery Rate (FDR) correction to adjust the p-values. All p-values reported in the main text were FDR-adjusted. All statistical analyses were conducted using R (v. 4.3.1, 2023).

## Results

### Sample description

Total of 80 patients (50 female) and 34 healthy controls (20 female) were considered for the final analysis (see Fig.1 for the patient selection steps). The percentage of male and female participants did not differ between the groups (χ²(1, N=114) = 0.03, p = 0.87). The median age of the recruited patients was 60 years (IQR = 13.25). Patients and healthy controls did not differ in age (U = 1227, p = 0.41). Male and female patients did not differ in this variable as well (U = 1495, p = 0.8). However, the statistically significant difference between groups in education years was identified (U = 1626, p = 0.013). There were also no differences in mean values of body mass index between patients and participants in the control group (t = –1.89, p = 0.06).

**Figure 1.**
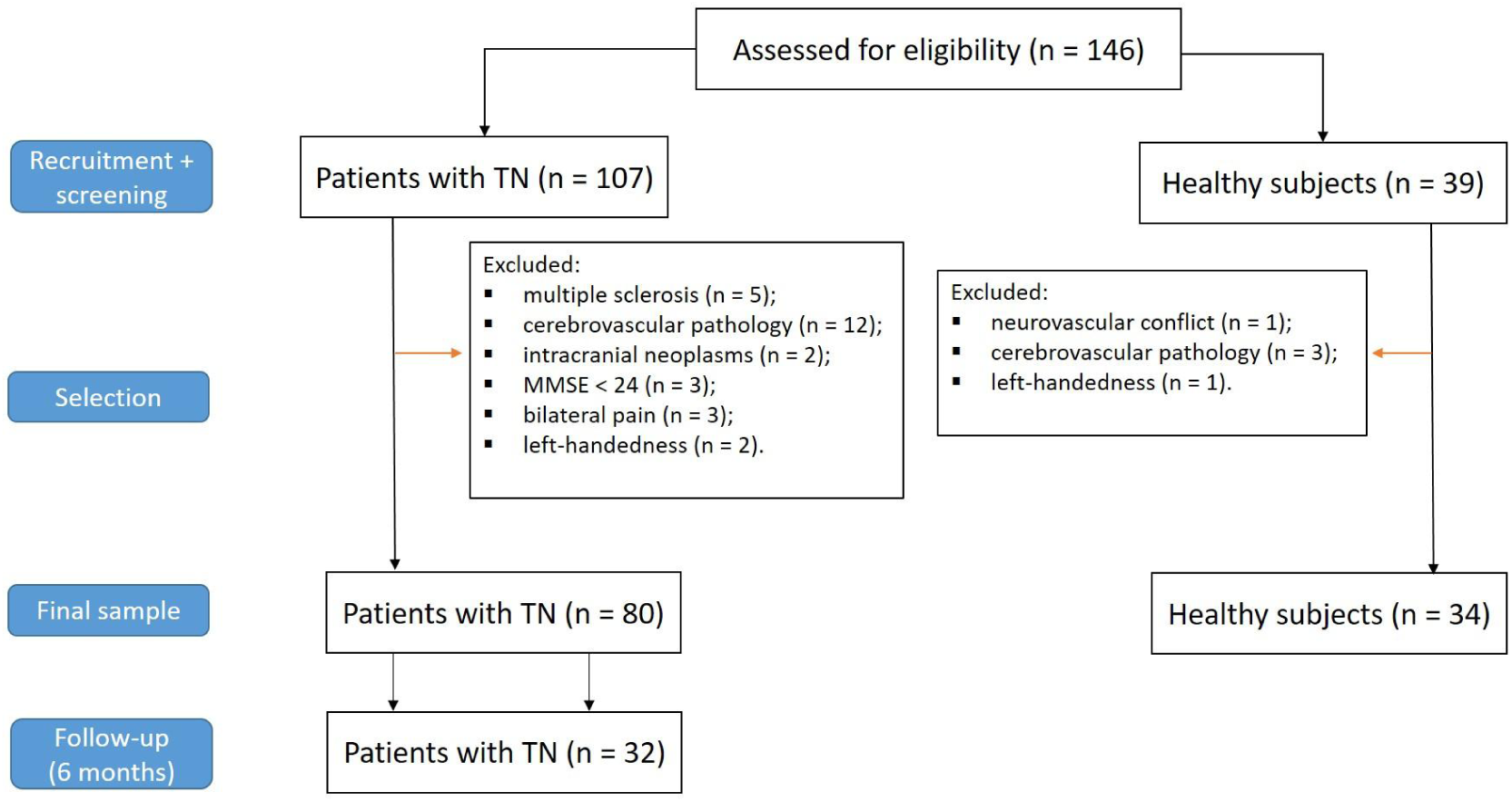
A flowchart illustrating the process of selecting patients based on study’s inclusion and exclusion criteria.

Forty eight patients (60%) had right-sided pain and thirty two patients (40%) suffered from left-sided pain. Median disease duration for the selected patients was 7 years (IQR = 7). Median value of BPI average scores were 5 (IQR = 2) and this value increased up to 7 (IQR = 4) for the BPI max scores. This data is summarized in Table 1. Hospitalized patients and those who received outpatient care did not differ in terms of pain severity (U = 183, p = 0.27) and disease duration (U = 219, p = 0.54). Before surgery, the most commonly taken drug was carbamazepine (85%), followed by gabapentin (9%), amitriptyline (7.5%) and pregabalin (6.3%). Detailed information on patients’ drug intake is provided in Supplemental Table 1. Seventy three patients from our sample underwent surgical intervention. Specifically, fifty five patients were subjected to microvascular decompression surgery (MVD; 75%), eleven patients underwent radiofrequency rhizotomy (RF; 15%) and balloon compression was performed on seven patients (BC; 10%). Patients with favorable and unfavorable surgical outcomes did not differ in terms of pain intensity (U = 354, p = 0.64) and disease duration (U = 415.5, p = 0.72). Eighteen out of 80 patients had a history of previous surgical interventions for facial pain (22.5%). Table 1 presents a concise overview of the key and most pertinent information regarding the patients’ main clinical variables.

**Table 1.**
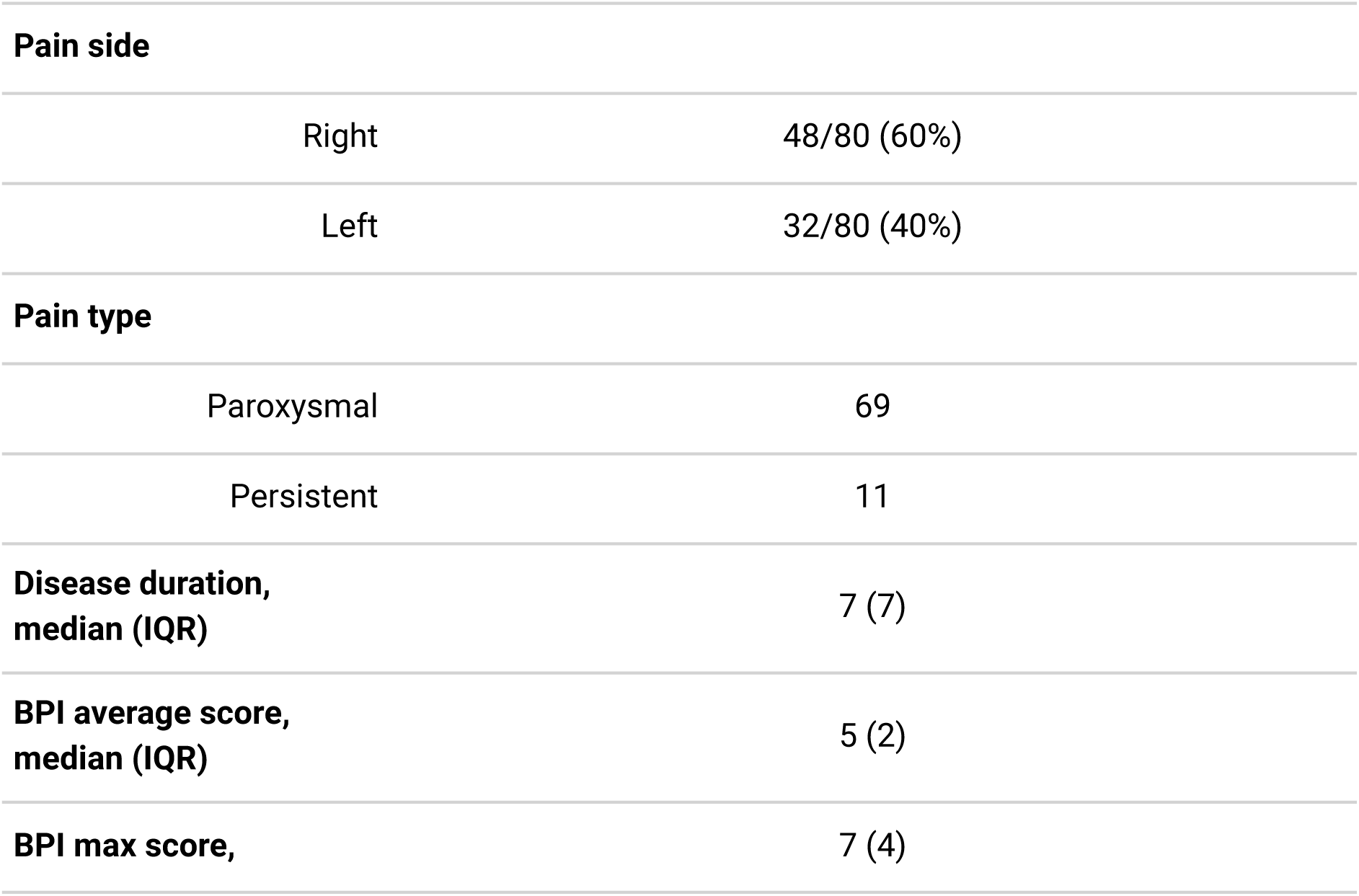
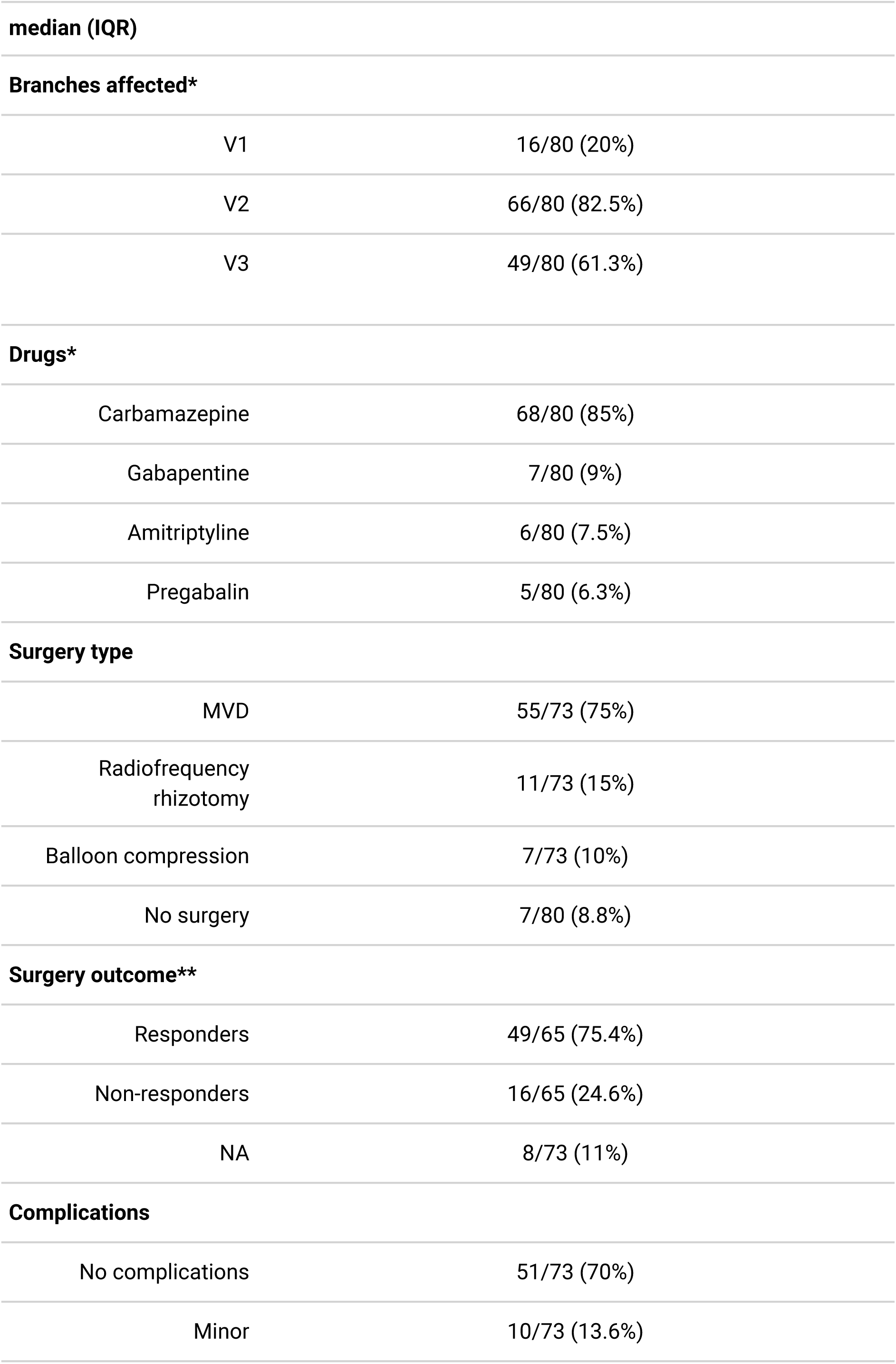
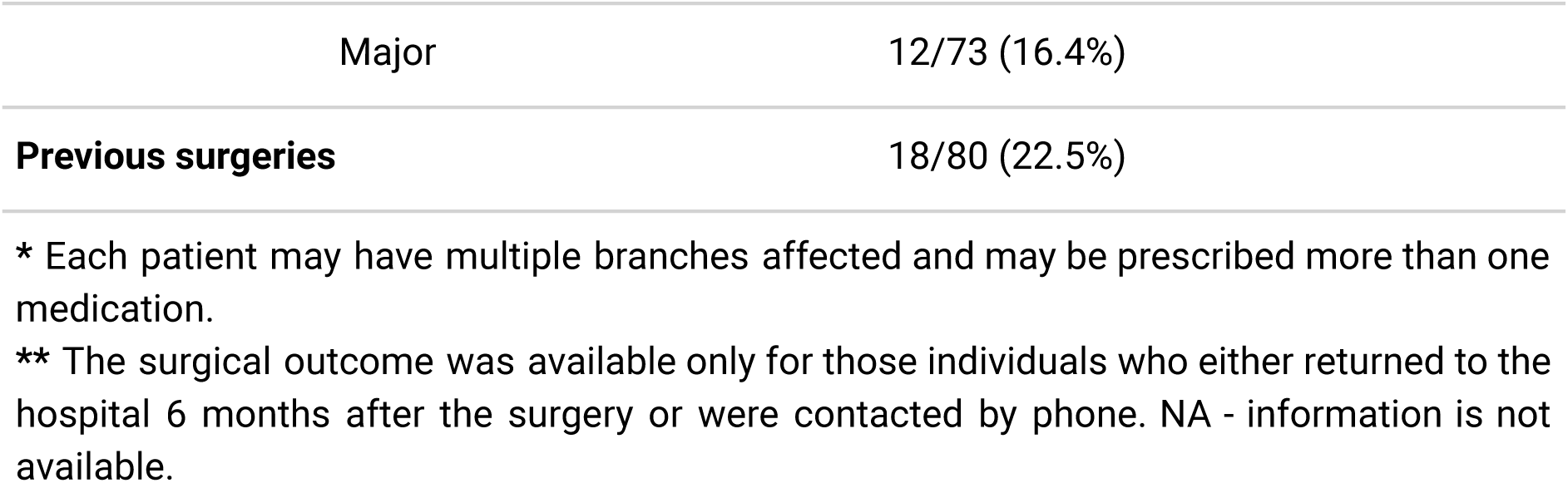
Clinical characteristics of TN patients.

### Neurocognitive testing

The distribution of neuropsychological scores on the tests used was non-normal, so a nonparametric Mann–Whitney test was employed to compare groups of healthy controls and patients with trigeminal neuralgia. Patients did not differ from healthy individuals in terms of MMSE scores (p = 0.42). None of the subtests in the ROCF test showed statistically significant differences between the groups either (all p > 0.1; Supplemental Table 2). The median scores of patients on the TMT and WAIS-F were at the same level as those of the control group (p > 0.1; Supplemental Table 2). At the same time, patients had lower scores on backward digit span test compared to healthy individuals (U = 1540, p = 0.024, r_rb_ = 0.31, 95% CI [0.08, 0.51]; Fig.2A). Phonemic and semantic verbal fluency scores were also diminished in patients with trigeminal neuralgia (U = 1797, p = 5.4*10^-5^, r_rb_ = 0.56, 95% CI [0.36, 0.70], Fig. 2B, and U = 1642, p = 0.003, r_rb_ = 0.42, 95% CI [0.20, 0.60], Fig. 2C, respectively).

**Figure 2.**
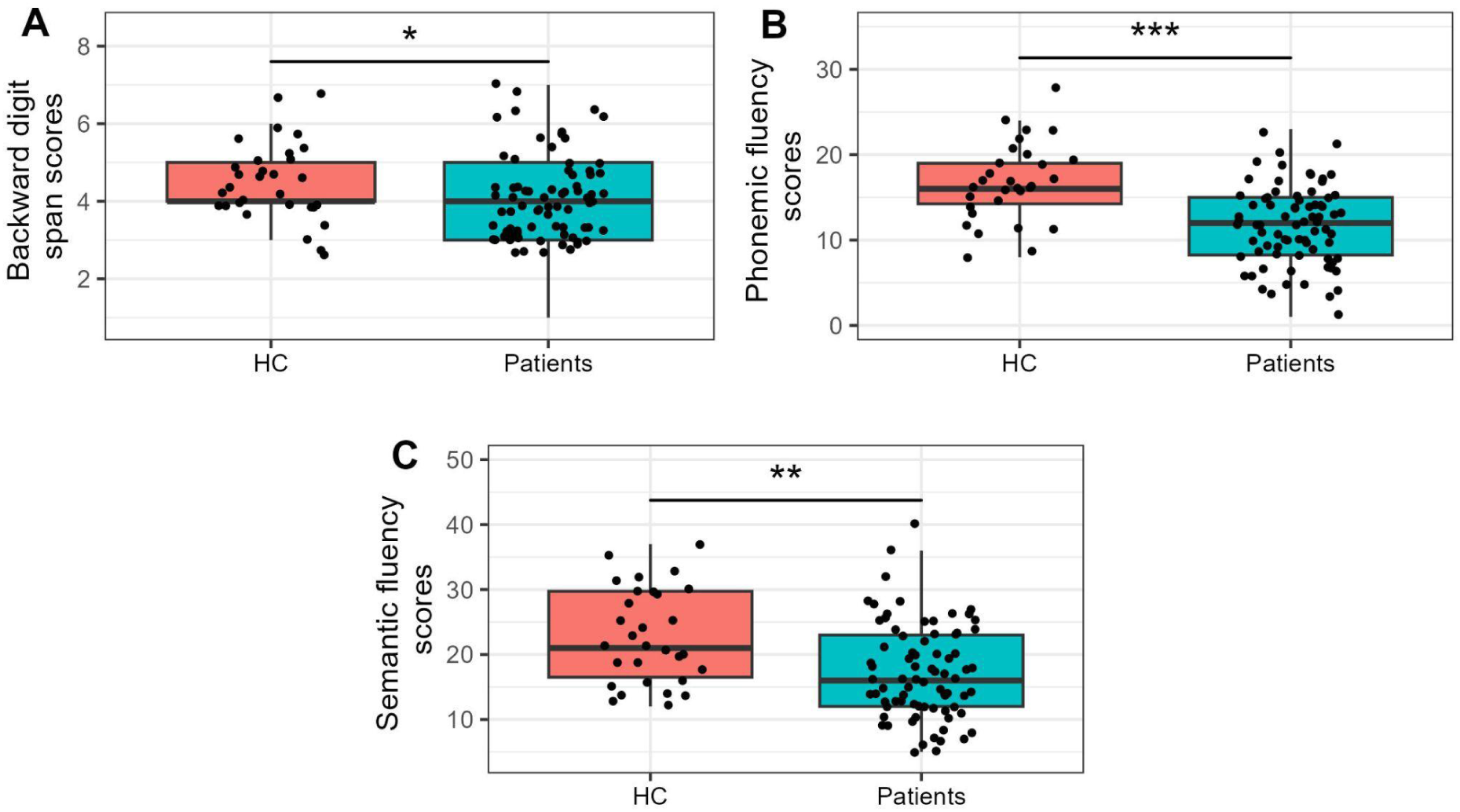
A visual representation of the differences in backward digit span (Panel A), phonemic (Panel B) and semantic fluency (Panel C) scores between control group and patients with TN. The asterisk signs indicate the level of statistical significance: * p < 0.05, ** p < 0.01, *** p < 0.001.

To ensure that the differences obtained were not influenced by potential confounding factors, we conducted an additional analysis using ANCOVA to control for depression and anxiety symptoms, pain catastrophizing, and educational attainment. ANCOVA is robust against mild to moderate violations of variables from the normal distribution, but it requires the model residuals to be normally distributed. We provide a detailed report on all the assumptions that need to be met for the model to be valid in Supplemental Figures 1-3. For WAIS-B scores, the main effect of the group was statistically significant after accounting for confounders (F(1,77) = 4.45, p = 0.038, η_p_^2^ = 0.05, 95% CI [0.002, 1]). In addition, we analyzed the possible confounding impact of attention (TMT-B part) and established that the differences in working memory scores between the groups are still evident (F(1,101) = 6.01, p = 0.016, η_p_^2^ = 0.06, 95% CI [0.006, 1]). Similarly, the results of comparing the scores on phonemic and semantic fluency tests remained statistically significant after additional analysis: F(1,76) = 14.85, p = 2.4*10^-4^, η_p_^2^ = 0.16, 95% CI [0.06, 1] for phonemic fluency and F(1,76) = 7.86, p = 0.006, η_p_^2^ = 0.09, 95% CI [0.02, 1] for semantic fluency task. We also examined the role of attention in influencing the differences in working memory between groups and found that these differences remained significant even after controlling for attention scores (Supplemental Table 3).

**Figure 3.**
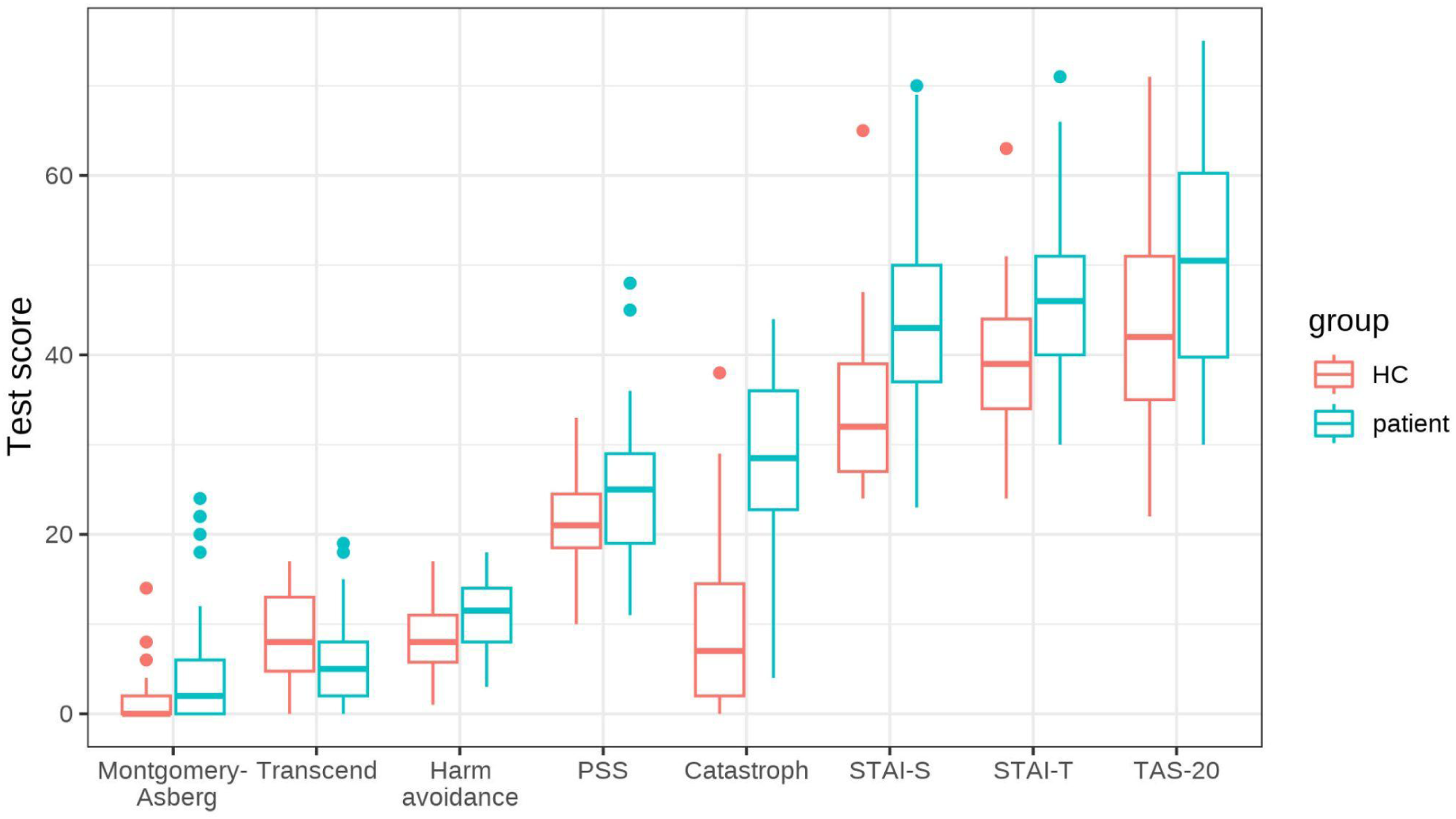
Grouped boxplots representing statistical comparison of scores of psychological tests and subscales between healthy participants and patients. Only the tests that showed statistically significant differences are presented here. Individual dots mark outliers — values of variables that fall outside the interval between Q_25_ – 1.5 IQR and Q_75_ + 1.5 IQR.

There were no differences in neurocognitive scores between patients with favorable and unfavorable postsurgical outcomes (all p > 0.1; Supplemental Table 4). Sensitivity analysis also demonstrated that excluding the outpatient individuals (n = 7) did not affect the results (Supplemental Table 5).

### Psychological assessment

The scores on the psychological tests and scales were normally distributed, except for the Montgomery–Asberg scale. For this scale, we used a nonparametric Mann–Whitney test to find the difference between groups of healthy individuals and patients. Otherwise, we employed a parametric t-test with Welch’s correction to account for unequal variances between the groups. Patients with TN showed significantly higher levels of state and trait anxiety compared to healthy controls (t = –5.1, p = 1.4*10^-5^, d = –1.06, 95% CI [-1.50, –0.62] and t = –3.76, p = 0.002, d = –0.82, 95% CI [-1.26, –0.37], respectively). Similarly, they had higher scores on the Perceived Stress Scale (t = –2.46, p = 0.02, d = –0.51, 95% CI [-0.93, –0.09]). When analyzing the subscales of the Cloninger Temperament and Character inventory, we found significant differences between groups for the Harm Avoidance and Transcendence subscales (t = –2.99, p = 0.01, d = –0.67, 95% CI [-1.12, –0.21] and t = 2.75, p = 0.02, d = 0.62, 95% CI [0.16, 1.07]). Patients with chronic pain also had elevated levels of depression (U = 642, p = 0.002, r_rb_ = –0.42, 95% CI [-0.60, –0.20]), pain catastrophizing (t = –8.38, p = 5.6*10^-11^, d = –1.81, 95% CI [-2.33, –1.29]) and alexithymia (t = –2.57, p = 0.02, d = –0.55, 95% CI [-0.98, –0.12]). A graphical representation of the comparisons is provided in Fig. 3.

Similarly to the neurocognitive scores, the results stayed the same after excluding outpatients without surgery (Supplemental Table 5). In addition, we repeated the entire analysis after excluding individuals with extreme score values. This sensitivity analysis showed that the removal of outliers did not affect the results in general, with the exception of the PSS variable, which lost its statistical significance (p = 0.07; Supplemental Table 6).

### Follow-up neuropsychological and psychological assessment

Data from thirty-two patients who were followed up for six months after surgery was available for analysis. Of the fifteen psychological and neuropsychological tests conducted, only three showed statistically significant differences between pre– and postsurgical conditions. There were no significant differences in neuropsychological scores between the two groups (p > 0.1). After adjusting for multiple comparisons, we observed notable contrasts in anxiety, depression, and pain catastrophization levels among the tested patients. The state anxiety scores, as measured using the Spielberger State and Trait Anxiety Questionnaire, were lower in patients six months after surgery compared to their presurgical levels (W = 146, p = 0.018, r_rb_ = 0.57, 95% CI [0.21, 0.81]; Fig. 4A). Similarly, the scores on the Montgomery–Asberg Depression and Pain Catastrophizing scales decreased following surgical treatment for trigeminal neuralgia (W = 120, p = 0.018, r_rb_ = 0.56, 95% CI [0.27, 0.76] and W = 202.5, p = 0.016, r_rb_ = 0.66, 95% CI [0.35, 0.85]; Fig. 4B and 4C respectively). Excluding patients who did not achieve complete pain relief after surgery (n = 7) did not affect the statistical significance of previously obtained results (Supplemental Table 7).

**Figure 4.**
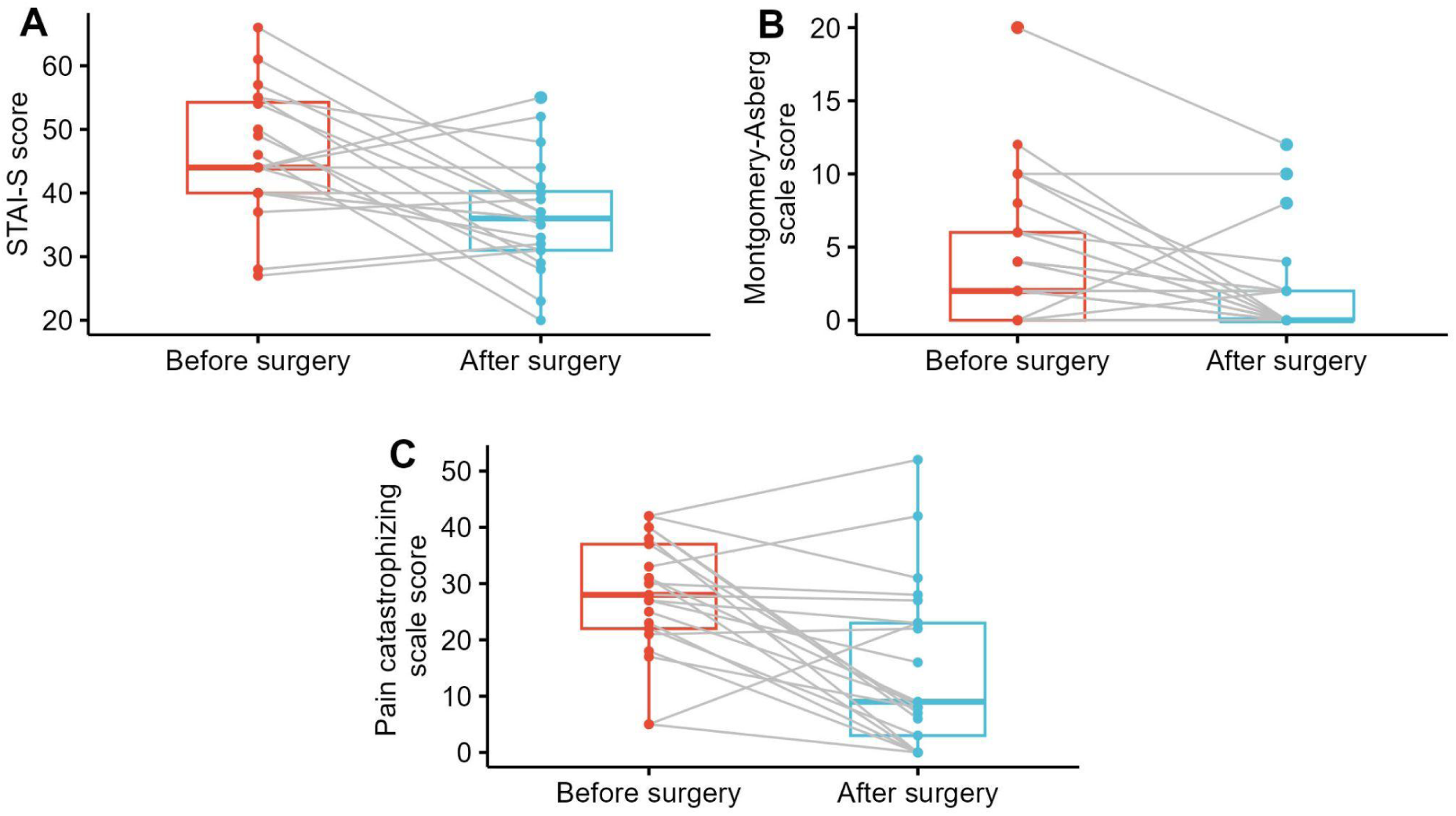
The results of comparing pre– and 6-month postoperative scores on the psychological tests, indicating statistical significance. Gray lines connecting the dots display the difference for individual patients.

### Correlation analysis results

Partial Spearman’s rho correlation was calculated between clinical and behavioral variables, taking into account the effect of age. A weak negative correlation was found between pain intensity and performance on the backward digit span test (rho = –0.29, p = 0.013). However, this result did not remain significant after correction for multiple comparisons, resulting in only a trend towards statistical significance (p = 0.08). Similarly, the same also applies to PSS scores, which were positively correlated with pain intensity (rho = 0.3, p = 0.019), but did not survive multiple comparison correction (p > 0.1). Moreover, we sought to estimate the impact of drug intake on neuropsychological performance scores, as it is well-established that antiepileptic drugs commonly prescribed to patients with TN can significantly affect cognitive functions. To this end, for the analysis we selected only patients who were undergoing carbamazepine monotherapy (n = 60). We did not find any significant associations between carbamazepine intake and cognitive function decline (all p > 0.1).

However, we were able to discover some significant associations between psychological and neuropsychological variables in patients with TN. Specifically, performance scores on ROCF delayed recall were negatively correlated with trait anxiety and depression symptoms, as well as alexithymia level (rho = –0.36, p = 0.046, 95% CI [-0.59, –0.08] for anxiety; rho = –0.38, p = 0.019, 95% CI [-0.58, –0.13] for depression and rho = –0.43, p = 0.019, 95% CI [-0.64, –0.17] for alexithymia scores). Beyond that, WAIS-F scores showed positive correlation with tolerance of uncertainty (rho = 0.4, p = 0.027, 95% CI [0.13, 0.61]) and negative correlation with alexithymia (rho = –0.38, p = 0.027, 95% CI [-0.59, –0.13]).

### Pain intensity prediction

To evaluate the ability of psychological and neurocognitive variables to predict pain intensity levels in patients with TN, we performed multiple linear regression. We conducted feature selection using four different approaches described in Section “Statistical analysis” to ensure the robustness and reliability of the overall procedure. We limited the maximal number of predictors included in the model to 5 in order to avoid overfitting. Based on the results of four independent feature selection procedures, we retained ROCF copy, phonemic verbal fluency, PSS and WAIS-B variables to be included in the final model. Following the removal of observations with missing values, the final linear model was built using data from 53 cases.

As shown in Table 2, three out of four selected predictors — namely, ROCF copy, phonemic verbal fluency and perceived stress score — were statistically significant, with the digit backward test variable approaching statistical significance (p = 0.06). The overall model with these predictors was statistically different from the null model (F(4, 48) = 6.42, p = 3.2*10^-4^). The multiple R-squared coefficient was equal to 0.35 and decreased to 0.29 after adjusting for the number of predictors. The LOOCV procedure provided an estimation of model errors after testing the model on new data: RMSE = 1.65 and MAE = 1.32. The verification of model assumptions is presented in Supplemental Figure 4.

**Table 2.** Final regression model for pain intensity scores (BPI average).

| Parameter | Coefficient | SE | t-value | p-value | 95% CI |
| --- | --- | --- | --- | --- | --- |
| Intercept | -2.82 | 2.25 | -1.25 | 0.22 | [-7.35, 1.71] |
| ROCF copy | 0.11 | 0.04 | 2.93 | 0.005 | [0.04, 0.19] |
| Verbal fluency (letter) | 0.14 | 0.05 | 2.55 | 0.014 | [0.03, 0.24] |
| PSS | 0.09 | 0.04 | 2.57 | 0.013 | [0.02, 0.16] |
| WAIS-B | -0.36 | 0.19 | -1.9 | 0.06 | [-0.75, 0.02] |

## Discussion

Chronic pain significantly affects the cognitive functions of patients suffering from various diseases characterized by persistent and severe pain, as demonstrated by noticeable decreases in performance scores on a variety of well-established and widely used neuropsychological assessments. Moreover, these patients may exhibit distinct and potentially unique combinations of personality traits and psychopathological symptoms. In this study, we conducted a comprehensive analysis of psychological and neuropsychological aspects of chronic pain in a relatively large group of patients with primary trigeminal neuralgia.

Our findings showed that the patients demonstrated the most significant decline in performance on phonemic and semantic fluency tasks, followed by those on backward digit span task. The first two tasks assess functioning of the semantic system, which is responsible for word storage and associations, as well as evaluating retrieval strategies, self-monitoring abilities, and inhibitory control of responses. The backward digit span task is a tool used to evaluate verbal working memory. A previous study conducted on a smaller sample of primary trigeminal neuralgia patients indicated that these individuals scored lower than the control group on composite memory, psychomotor speed, reaction time, complex attention, and cognitive flexibility tasks (Meskal, 2014). However, in our study, no significant differences were observed between the groups with regard to attention and short-term and long-term non-verbal (visuospatial) memory functioning. The same was true for cognitive flexibility scores, as measured with Trail Making test part B in this study, where we found no differences. This finding is consistent with the results of a recent study by Howlett and colleagues, which argues against general impairment in cognitive flexibility in patients with chronic pain (Howlett et al., 2024). It should also be noted that currently there is still no consensus in this research area, given that findings of other studies frequently point to the opposite direction. For example, median TMT scores did not differ significantly between the groups, which was previously demonstrated for patients with various chronic pain conditions (Tesio et al., 2015; van der Leeuw et al., 2018).

Nevertheless, in this study we were able to replicate impairments in working memory, especially in tasks with a high memory load, as demonstrated by the backward part of the digit span test. These findings further strengthen and support the available evidence for transdiagnostic impairment of executive functions in individuals with chronic pain, although specific domains affected may vary across different conditions (Berryman, 2014). Prior studies have consistently shown that patients experiencing chronic pain exhibit reduced working memory scores (Berryman, 2013; Baker, 2016; Mazza et al., 2018). Moreover, it is recognized that working memory capacity is associated with pain intensity, such that pain perception tends to be lower during tasks with high working memory demands compared to less cognitively demanding conditions (Nakae et al., 2013; Torta et al., 2020; Deldar et al., 2021). The hypothesis of limited cognitive resources has been proposed to explain empirical findings indicating that pain processing and cognitive tasks engage overlapping neural circuits, potentially competing for access to executive functions. In situations of heightened nociceptive input, the salience and behavioral importance of pain stimuli may dominate, diverting resources from complex cognitive processing. Another plausible explanation for these results is that chronic intense pain could be viewed as a persistent chronic stressor, a well-documented factor linked to cognitive decline and gray matter reduction in the prefrontal cortex (Abdallah & Geha, 2017; Jacobsen et al., 2023). Notably, some authors have previously suggested and empirically demonstrated that the observed differences in working memory scores may stem from diminished attentional capacity (Oosterman et al., 2011; Oosterman et al., 2012). However, after controlling for the potential mediating role of attention, we found that there were still significant differences in working memory performance between the groups.

Cognitive decline in individuals with chronic pain could potentially exacerbate their conditions over time, either by amplifying pain perception or diminishing the efficacy of pain-relieving medications (Kunz et al., 2015). The cognitive task that revealed the most pronounced group disparities was the verbal fluency test, which assesses lexical access and executive control abilities. While the differences observed in this task may be attributed to a decline in the executive control component, similar to the working memory test, an alternative explanation could be more closely linked to lexical access and semantic memory systems. Building on previous research, it has been suggested that persistent pain might give rise to the formation of multiple «pain memories», associating instances of pain with otherwise benign environmental contexts and internal states (Phelps et al., 2021). Current treatments for chronic pain could potentially work by extinguishing these pain-related memories, alongside or instead of their effects on sensory systems. While speculative, deficits in semantic and possibly episodic memory retrieval and control could serve as a compensatory mechanism aimed at shielding the self from recalling negatively charged memories (Swannell et al., 2016). This, in turn, might contribute to the onset of depression or depression-like behaviors, as indicated by findings from several clinical studies (Nekovarova et al., 2014).

In general, while being consistent with existing literature findings regarding the decline in neurocognitive performance scores across different domains, it is important to be cautious when interpreting these results in terms of specific affected executive functions. Different research groups may attribute the observed neurocognitive deficits to various executive functions or core executive domains such as inhibition, shifting, updating, or access, which can arise from the so-called task-impurity problem — the inability of a specific task to consistently tap into one particular latent construct that this task is intended to measure (Ambrosini et al., 2019). Nevertheless, upon independent validation, these findings could inform the development of targeted cognitive interventions aimed at the most affected domains in individuals with chronic pain more broadly, and specifically in those with trigeminal neuralgia, to alleviate pain severity and enhance the quality of life for patients.

It is worth noting that in this study, we also sought to assess the impact of carbamazepine intake on cognitive functions. Carbamazepine is known to have some adverse side effects, such as psychomotor retardation, dizziness, and excessive sleepiness, which may lead patients to discontinue therapy (Ashina et al., 2024). However, our findings did not suggest a correlation between the carbamazepine dose and a decline in cognitive functions. Patients with chronic trigeminal pain often attribute their decreased performance on neuropsychological tests to the influence of their medication, although this has not yet been conclusively proven, given conflicting published findings (Delcker et al., 1997; Tentolouris-Piperas et al., 2018).

In addition to diminished neurocognitive performance scores, individuals with chronic trigeminal neuralgia demonstrated heightened state and trait anxiety levels, along with increased perceived stress and depressive symptoms. These findings are in line with the extensive body of literature highlighting the common co-occurrence of pain with depression and anxiety, including the results of our recent investigation (Moisak et al., 2021). Moreover, there is evidence suggesting that certain psychopathological symptoms and personality traits may influence the relationship between pain intensity and cognitive functioning measures (Castel et al., 2021). Beyond that, research conducted by Apkarian and colleagues over the past decade has compellingly demonstrated a shift in brain activity among chronic pain patients, transitioning from primarily sensorimotor regions to limbic areas as the condition progresses. This shift may imply an increasing significance of emotional factors in the maintenance of pain during the chronic phase of the disease (Baliki and Apkarian, 2015).

Our patient cohort also showed differences compared to the control group in their ability to recognize and express emotions. Elevated alexithymia scores are often observed in individuals with various chronic pain conditions and are associated with increased pain severity and functional impairment (Aaron et al., 2019; Asgarabad et al., 2023). In relation to facial pain, recent reports indicate that alexithymia may serve as a prognostic factor for surgical outcomes in patients with trigeminal neuralgia (Samanci et al., 2023). Another line of evidence highlights the strong connections between alexithymia and cognitive functioning. Individuals with higher alexithymia scores tend to exhibit poorer memory, less effective emotion regulation strategies, and impaired performance on verbal executive function tasks (Santorelli and Ready, 2015; Correro et al., 2021; Preece et al., 2023). In this study, we identified moderate negative correlations between alexithymia and memory performance scores. Specifically, elevated levels of alexithymia were associated with lower scores on the ROCF delayed recall and forward digit span tasks. Notably, these associations with cognitive functioning were found to be stronger than those related to anxiety and depression, which are well-established contributors to executive dysfunction across various disorders, regardless of the presence of chronic pain. Since alexithymia is thought to reflect an underlying interoceptive deficit, and it has been empirically demonstrated that interoceptive sensitivity directly influences pain perception, it would be advantageous to further explore the extent of interoceptive failures in patients with trigeminal neuralgia and their therapeutic applications in future research (Di Lernia et al., 2016).

Similarly, our study revealed intergroup variations in pain catastrophizing scores, suggesting that patients with a prolonged history of pain held more pronounced negative beliefs associated with pain compared to control participants. Pain catastrophizing is a well-studied psychological phenomenon and cognitive vulnerability factor characterized by excessive worrying about pain, an inability to control fear associated with pain and feeling powerless to cope with pain. Prior studies have consistently linked pain catastrophizing to increased likelihood of developing chronic pain after surgery, pain persistence and poorer quality of life (Khan et al., 2011; Reiter et al., 2018). Interestingly, experimental manipulation of pain catastrophizing levels in both chronic pain patients and healthy subjects had a significant impact on experienced pain intensity (Kjøgx et al., 2016). The accumulated body of evidence suggests that this trait may be a modifiable factor of pain severity and is now widely regarded as a valid and promising target for treatment in psychotherapy for patients with chronic pain (Schütze et al., 2018; Gilliam et al., 2021). In general, our findings are in full accordance with previous experimental results reporting increased pain catastrophizing levels among patients with trigeminal neuralgia (Zakrzewska et al., 2017).

Lastly, using the Temperament and Character Inventory, we found that individuals with trigeminal neuralgia scored higher on the harm avoidance scale. It is worth noting that this observation is not unique to trigeminal neuralgia but can be seen across different chronic pain syndromes (Knaster et al., 2012; Leombruni et al., 2016). The harm avoidance trait has been linked to increased pain sensitivity and less effective conditioned pain modulation responses in healthy individuals subjected to cold pressor tests and thermal paradigms for conditioned pain modulation (Pud et al., 2004; Nahman-Averbuch et al., 2016). Interestingly, previous work by Tuominen and colleagues showed that an elevated harm avoidance score correlated with increased availability of μ-opioid receptors (indicative of reduced endogenous μ-opioid activity) in frontal regions involved in emotion and pain regulation (Tuominen et al., 2012). Further research is needed to determine whether addressing the harm avoidance trait could serve as a viable therapeutic approach in innovative psychological interventions tailored for individuals with chronic pain. The second subscale on which patients differed from healthy controls was a Self-transcendence subscale. By evaluating responders’ attitudes towards spirituality, religion and supernatural phenomena, this tool can offer valuable insights into a patient’s personality. Higher scores on this scale are shown to positively correlate with psychotic traits both in clinical and healthy samples (Nitzburg et al., 2020). Some authors suggested that self-transcendence may serve as a protective factor or coping strategy, mitigating the negative consequences of depressive mood, ruminations, and excessive worrying (Josefsson et al., 2011; Schwalm et al., 2022). It is important to note that, in contrast to patients with fibromyalgia, who scored higher than the control group subjects on this scale, we found that patients with trigeminal pain showed lower self-transcendence compared to healthy subjects (Lundberg et al., 2009; Gencay-Can and Suleyman Can, 2012). From this perspective, the low level of self-transcendence trait in our patient sample may reflect a downregulation of stress-coping adaptive mechanisms.

In this study, we also demonstrated that variables derived from neuropsychological and psychological assessments can effectively predict pain intensity. Specifically, measures of visuospatial abilities, working memory and semantic fluency, along with perceived stress scores, were identified as significant predictors in our model, explaining 29% of the variance in pain intensity. These results are consistent with previous studies suggesting that psychological variables can reliably predict a wide range of pain characteristics, including intensity, duration, and unpleasantness (Bainter et al., 2020). ROCF copy scores, although not statistically different between groups, were nevertheless a significant contributor to the variance of dependent variable explained by the model. In earlier studies, Lee and colleagues also failed to detect any differences in ROFC copy scores between patients with widespread chronic pain and subjects in the control group (Lee et al., 2010). However, unlike our findings, they did not establish a relationship between pain status and ROCF-copy scores. Interestingly, Galvez-Sanchez and coauthors were able to find differences in ROCF performance scores between experimental and control groups on a sample of fibromyalgia patients (Galvez-Sanchez et al., 2018). One possible explanation for these discrepancies may stem from the fact that different chronic pain conditions can affect cognitive domains differently. Another possible reason could be due to the differences in scoring systems used to rate patients’ performance on this test. While Lee and Galvez-Sanchez groups used partition into 18 elementary parts with two points being ascribed for accurate reproduction of the particular part, resulting in a total of 36 points, we employed division into 48 elements (48 points) proposed by Petilli et al., 2021. At the same time, Attal and colleagues followed up a large sample of patients who had undergone total knee arthroplasty or breast cancer surgery at 6 and 12 months after the surgery in order to identify neuropsychological predictors of pain development later on (Attal et al., 2014). They demonstrated that performance on the TMT-B, ROCF copy, as well as ROCF recall were predictive of the presence of pain at these time points. Taken together, the heterogeneity of samples and research designs employed precludes drawing definitive conclusions based on the cited publications, necessitating further high-quality studies.

It is also worthwhile to mention several limitations of this study. First, the current study design does not allow us to confidently determine whether chronic pain is the main and primary cause of cognitive decline. Instead, pre-existing cognitive impairments and personality traits may make some individuals more likely to develop chronic pain following an acute episode. Second, the outcome of the surgery was determined by the presence or absence of pain six months later. Some authors used the assessment of patients’ pain status immediately after surgery or even a year later as the outcome, which can significantly affect the results ultimately obtained. At the same time, this could also influence the lack of significant differences in neuropsychological tests six months later. In future studies, it would be informative to evaluate the neuropsychological status of patients one year after the surgery. Third, the sample size of patients in the follow-up study was small (n = 32), which could also have influenced the results of the study. Fourth, in this study, we did not find statistically significant differences between the groups in terms of cognitive flexibility using TMT-B. To confirm the data obtained, it is advisable to use other tools for assessing cognitive flexibility, such as the Stroop test, Wisconsin Card Sorting Test or Towers of London. Fifth, we cannot rule out the possibility that statistically significant changes on 3 psychological scales (STAI-S, Montgomery–Åsberg scale and pain catastrophizing scale) when comparing before and after surgery are not related to an increased level of anxiety before surgery, but are a direct consequence of the pain relief. Future studies may be enlightening in addressing this issue, explicitly taking into account patients’ apprehension about impending surgery. Sixth, we did not find an effect of carbamazepine dosage on cognitive functions. It should be noted, however, that many patients had previously taken other medications, and their influence on the patient’s current cognitive status is difficult to assess. Also, depending on their pain level, patients changed the dosage of the medication they were taking. Finally, all neuropsychological tests were evaluated by only one neuropsychologist, which can cause some problems. For example, evaluation of ROCF test scores can vary significantly between different specialists, so in future studies it would be desirable to have an independent evaluation by two neuropsychologists with calculation of the inter-rater agreement index.

In general, the conclusions of the article should be interpreted with caution, since it is currently unknown whether they can be generalized to all patients with trigeminal neuralgia, or whether they are specific only to patients with chronic, intense pain for whom pharmacotherapy loses its effectiveness over time.

## Author contributions

Pashkov A. – methodology, investigation, formal analysis, visualization, writing – original draft; Filimonova E. – investigation, data curation, writing – review & editing; Martirosyan A. – investigation; Moisak G. – conceptualization, writing – review & editing, supervision; Rzaev J. – conceptualization, writing – review & editing, supervision.

## Supplemental Materials

## Data Availability

All data produced in the present study are available upon reasonable request to the authors

